# Excess weight mediates changes in HDL pool that reduce cholesterol efflux capacity and increase antioxidant activity

**DOI:** 10.1101/19002899

**Authors:** Jose Carlos de Lima-Junior, Vitor W.M. Virginio, Filipe A. Moura, Adriana Bertolami, Marcelo Bertolami, Otavio R Coelho-Filho, Ilaria Zanotti, Wilson Nadruz, Eliana Cotta de Faria, Luiz Sergio F. de Carvalho, Andrei C Sposito

## Abstract

**Objective:** Obesity-related decline in high-density lipoprotein (HDL) functions such as cholesterol efflux capacity (CEC) has supported the notion that this lipoprotein dysfunction may contribute for atherogenesis among obese patients. Besides, potentially other HDL protective actions may be affected with weight gain and these changes may occur even before the obesity range.

**Methods:** Lipid profile, body mass index (BMI), biochemical measurements, and carotid intima-media thickness (cIMT) were obtained in this cross-sectional study with 899 asymptomatic individuals. HDL functions were measured in a subgroup (n=101).

**Results:** Individuals with increased HDL-C had an attenuated increase in cIMT with elevation of BMI. CEC, HDL-C, HDL size and HDL-antioxidant activity were negatively associated with cIMT. BMI was inversely associated with HDL-mediated inhibition of platelet aggregation and CEC, but surprisingly it was directly associated with the antioxidant activity. Thus, even in non-obese, non-diabetic individuals, increased BMI is associated with a wide change in protective functions of HDL, reducing CEC and increasing antioxidant activity. In these subjects, decreased HDL concentration, size or function are related to increased atherosclerotic burden.

**Conclusion:** Our findings demonstrate that in non-obese, non-diabetic individuals, the increasing values of BMI are associated with impaired protective functions of HDL and concomitant increase in atherosclerotic burden.

## 1. Introduction

Presently, one in three individuals with excess weight will die from cardiovascular disease (CVD)[1, 2]. In fact, both obesity (BMI >30 kg/m^2^) and overweight (BMI of 25 to <30 kg/m^2^) individuals are at increased risk of cardiovascular death as compared with those with BMI within the normal range (18.5 to <25 kg/m^2^)[3]. The proposed milieu for this interaction involves a spectrum of mechanisms in which phenotypic or functional changes in high-density lipoprotein (HDL) are involved [4].

The identification of HDL involvement in adiposopathy is not a recent issue. Since the metabolic syndrome was conceived, all clinical criteria for its diagnosis included low levels of plasma HDL-cholesterol (HDL-C) as a marker of metabolically, unhealthy obesity. In the mechanistic point of view, the combination of increased substrate, such as triglyceride-rich lipoproteins, and increased activity of HDL remodeling proteins, such as cholesteryl ester transfer protein (CETP), and hepatic lipase (HL), coexists in overweight individuals as a result of insulin resistance, promoting a reduction in HDL concentration, size and its content of apolipoproteins AI (ApoA-I). In addition, a decrease in the overall cholesterol efflux capacity (CEC) has been reported in obese subjects [5], despite the increased ABCA1-mediated cholesterol efflux[6].

Besides CEC, HDL also mediates several other anti-atherosclerotic mechanisms, such as antioxidant, anti-inflammatory activities and inhibition of platelet aggregation, whose extent is sensitive to phenotypic changes of the particles, such as those described above. If this is so, more than the reduced capacity as free cholesterol acceptor, an overall dysfunction in the HDL system may follow weight gain [7]. Furthermore, as the magnitude of these phenotypic changes in HDL occurs in parallel with the increase in BMI, it is possible that a cluster of particle dysfunctions occurs earlier with the weight gain, possibly even before the criterion for overweight. Hence, a metabolic legacy related to HDL may contribute to the future CVD risk. In order to shed some light on these gaps, we designed this study to evaluate the impact of BMI on the interaction between HDL concentration and functions with atherosclerotic burden in pre-obese individuals.

## 2. Materials and methods

### 2.1. Cross-sectional study description

We evaluated a sample of 899 asymptomatic individuals who were seen in two primary care centers between 2008 and 2013: the outpatient clinic at Dante Pazzanese Institute of Cardiology, São Paulo (SP), Brazil, and governmental primary care centers of the city of Campinas, SP, Brazil. Inclusion criteria were: (1) no manifested atherosclerotic CVD; (2) no diagnosis of type 2 diabetes based on antidiabetic treatment, fast blood glycemia ≥126 mg/dL, glycated hemoglobin ≥6.5% or glycemia ≥200 mg/dL on 120-minutes oral tolerance test and (3) age between 20 and 75 years old. We excluded individuals with (1) uncontrolled hyper (thyroid stimulating hormone (TSH) <0.41μUI/mL or free thyroxin >1.8ng/dL) or hypothyroidism (TSH>4.50μUI/mL or free thyroxin <0.9 ng/dL); (2) antidiabetic medications; (3) liver disease, as indicated by ALT or AST over two times the upper limit; (4) urea >40mg/dL; (5) glomerular filtration rate ≤60 mL/min/1.73m^2^; (6) heart failure NYHA stage ≥III; (7) HIV positive or (8) withdrawal of informed consent. Patients underwent clinical examination, as well as biochemical analysis. Subclinical atherosclerosis was measured in all patients up to a month after being included. The study was approved by the local institutional ethical committee of the Instituto Dante Pazzanese de Cardiologia (registration number 3852/2009). It is also registered at ClinicalTrials.Gov by the identification NCT02487615. All patients provided written and informed consent forms before taking part in the study. The study protocol conforms to the ethical guidelines of the 1975 Declaration of Helsinki.

A sub-analysis testing functions and characteristics of HDL was performed in consecutive sample of 101 individuals from the enrolled participants in the governmental primary care centers of the city of Campinas. The study was approved by the local institutional ethical committee of the Hospital das Clínicas of the State University of Campinas (registration number 1260/2010). It is registered at ClinicalTrials.Gov by the identification NCT02106013. All patients provided written and informed consent forms before taking part in the study. The study protocol conforms to the ethical guidelines of the 1975 Declaration of Helsinki.

### 2.2. Biochemical analysis

Blood samples were drawn after a 12-hour fasting period. The following biochemical measurements were performed: triglycerides, total cholesterol, HDL-C, creatinine, c-reactive protein (CRP), insulin, glucose, and insulin. LDL-C was calculated using Friedewald’s equation. Glomerular filtration rate was calculated by the CKD-EPI equation. HOMA2 % S, HOMA2-IR and HOMA2%β were calculated using computer models [8].

### 2.3. Carotid artery ultrasound

Carotid Doppler ultrasound was performed using high-resolution Vivid 7 ultrasound (GE, USA) and high-frequency linear transducer (9 MHz) with automatic border recognizer detection as described previously in Bertolami et al [9]. Briefly, the cIMT was obtained by means of image processing of B-mode ultrasonograms of the right and left automatic measurement. High-resolution B-mode ultra-sonographic imaging was performed initially evaluating the common carotid artery with antero-oblique insonation above the clavicle and alongside the internal carotid artery, as standardized procedure[10]. Measurement of cIMT was obtained 20 mm proximally from the carotid bifurcation as the distance between the lumen– intima interface and the media–adventitia interface[11].

### 2.4. Lipoprotein isolation

LDL was isolated from a pool of normolipidemic sera from 20 volunteers, through sequential ultracentrifugation using a Beckman L8-M ultracentrifuge (Beckman Coulter Inc., Palo Alto, USA), with a 75Ti fixed angle rotor (Havel, 1955 #11). HDL was isolated from each study participant through density gradient ultracentrifugation[12] with the use of a SW41Ti rotor. Isolated lipoproteins were extensively dialyzed against EDTA-free PBS for 24h, at 4^°^C, in a dark room. All assays were performed in freshly isolated lipoproteins that were kept at 4^°^C for a maximum period of 15 days.

### 2.5. HDL chemical composition and molar concentration measurements

HDL chemical composition was measured using commercially available enzymatic kits, in the microplate reader Power Wave XS (BioTek®, Winooski, USA). Total proteins (Pierce(tm) BCA Protein Assay Kit, Thermo Scientific, Rockford, USA), TC (CHOD-PAP, total cholesterol, Roche Diagnostics® reagents, Mannheim, Germany), FC (Free Cholesterol E, Wako Chemicals, Richmond, USA), PL (Phospholipids C, Wako Chemicals, Richmond, USA), TG (TG, GPO-PAP, Roche Diagnostics® reagents, Mannheim, Germany) and ApoA-I (TINA QUANT APOA1 V2, Roche Diagnostics® reagents, Mannheim, Germany) were measured, while CE was calculated according to the following formula: (TC–FC) x 1.67[12]. The relative content of ApoA-I (HDL-ApoA-I) or lipids in HDL was calculated based on their proportion to the total mass of HDL, calculated as the sum of FC, PL, TG, CE, and total proteins. HDL molar concentration was estimated based on particle total mass and molecular weight[12].

### 2.6. HDL physical-chemical characterization

HDL particle size was determined using dynamic light scattering, in a Nanotrac Particle Size Analyser 250 (Microtrac Inc., Montgomeryville, USA)[13]. Zeta potential was determined in HDL diluted 1:10 in KCl 10mM, using laser Doppler micro-electrophoresis, in the Zetasizer Nano ZS (Malvern Instruments, Malvern, UK).

### 2.7. Determination of proteins involved in HDL metabolism

CETP and PLTP activities were measured using exogenous radiometric assays, as previously described[14, 15]. LPL and HL activities were measured in fasted post-heparin plasma samples, collected 15min after the intravenous administration of heparin (100U/kg body weight), in an assay based on fatty acid release from a radiolabeled triolein emulsion[16]. LCAT activity was determined using recombinant HDL, according to standardized method [17]. PON activity was measured using paraoxon (diethyl-p-nitrophenylphosphate, Sigma, St. Louis, MO, USA) as substrate [18].

### 2.8. HDL antioxidant activity

HDL antioxidant activity was measured in a kinetic fluorimetric assay adapted from Navab et al[19]. Oxidation was monitored as changes in the fluorescence intensity of 2’,7’-dichlorofluoresceine (DCFH). For the antioxidant activity assays, LDL (final concentration, 20mgTC/dL) and CuSO_4_ (final concentration, 0.5μM) were added to DCFH-containing tubes (final concentration, 2mg/mL), followed by the addition or not of HDL (final concentration, 15mg total mass/dL). The volume was adjusted to 100µL with Chelex treated-PBS and the reaction mixture transferred onto a black 96-well microplate. The plate was covered with an optical adhesive cover to avoid evaporation and incubated at 37°C. Fluorescence intensity was measured over 24h with 15-minute intervals in a fluorescence microplate reader (Spectra Max M5; Molecular Devices, Sunnyvale, USA) at an excitation wavelength of 485nm, emission wavelength of 540 nm, and cut-off of 530nm. Results of antioxidant activity are presented as the percentage of inhibition of LDL oxidation in the presence of each subject’s HDL when compared to control wells (LDL alone).

### 2.9. HDL anti-inflammatory activity

HDL’s anti-inflammatory activity was measured in HUVEC in an assay adapted from Besler et al[20]. Cells were cultured in RPMI 1640 medium containing 10% fetal calf serum, penicillin, and streptomycin and maintained in a 5% CO_2_ incubator at 37^°^C. After reaching confluence, they were plated in 24-well culture plate (3 ⨯ 10^5^ cells/well, and incubated with TNF-α (1ng/mL), with or without HDL (50μg ApoA-I/mL) for three hours. The culture media were collected and stored at -80^°^C. Due to the low VCAM-1 concentrations, samples were concentrated using the Amicon® Ultra Centrifugal Filters, 50K (Millipore, Massachusetts, USA) and then VCAM-1 concentrations were measured using the Human VCAM-1 ELISA Kit (Cat number ECM340, Millipore, Massachusetts, USA). Results are expressed as the percentage of decrease in VCAM-1 concentrations in the wells incubated with HDL when compared to the control wells without HDL.

### 2.10. Cholesterol efflux capacity (CEC) assay

Global cellular CEC was performed using J774 macrophages enriched with acetylated LDL and ^14^C-cholesterol and HDL as the cholesterol acceptor[21]. In summary, J774 macrophages were cultured in RPMI 1640 medium containing 10% fetal calf serum, penicillin, and streptomycin and maintained in a 5% CO_2_ incubator at 37°C. After reaching confluence, cells were plated in a 96-well plate (1.25 ° 10^5^ cells/well, and enriched with acetylated LDL (50 μg/mL) and ^14^C-free cholesterol (0.3μCi / mL). After 48h, cells were washed with PBS containing fatty acid-free albumin (FAFA) and equilibrated with DMEM containing FAFA for 24 hours. The cells were then washed twice and incubated with HDL (50μg ApoA-I/mL) for 8 hours. Media were collected and the radioactivity measured in a beta-scintillation counter. Cells were rinsed twice with cold physiologic saline and the intracellular lipids extracted with hexane: isopropanol (3:2, v/v). Solvent was evaporated and radioactivity measured. The percentage of ^14^C-CEC was calculated as (^14^C-cholesterol in the medium/^14^C-cholesterol in cells+medium) ×100.

### 2.11. HDL-mediated platelet aggregation inhibition

HDL ability to inhibit platelet aggregation was measured as described by Valiyaveettil et al[22]. HDL (0.8mg protein/mL) was mildly oxidized by dialysis against PBS + 5μM CuSO_4_, for 24h at 37°C. Citrated blood was drawn from a healthy donor and platelet-rich plasma (PRP) obtained by centrifugation at 800rpm for 15 minutes. PRP was then incubated with native or oxidized HDL (0.5 mg/mL) for 30 minutes at 37°C. Platelet aggregation was monitored using a Lumi-Aggregometer type 500 VS (Chrono-log, Havertown, USA) for 6 minutes after the addition of ADP (5μM). Results are expressed as percentage of the inhibition of platelet aggregation induced by oxidized compared to native HDL.

### 2.12. Statistical analysis

The distribution of all variables was tested with Kolmogorov-Smirnov test. Jonckheere-Terpstra trend in one-tail test was used to evaluate demographic, clinical variables and HDL characterization among three groups clustered using BMI and expressed as median and interquartile range. ANCOVA was used to perform comparisons among groups involving enzymatic components of HDL adjusted for age, sex, and HOMA2 %S. Generalized linear models with type III sums of squares were used to assess interactions of BMI within the association between HDL functions and cIMT, the only outcome, adjusted for age, sex, HDL-C, BMI, and HOMA2 %S. HDL functions and HDL-C were modeled with generalized linear regression. The BMI effect on the cIMT among six HDL functions (HDL-C, HDL size, antioxidant activity, CEC, HDL-mediated platelet inhibition, and anti-inflammatory activity) was evaluated with the use of interaction tests. We used complete case analysis to handle missing data. To construct the 3D surface plot 1, we included only HDL-C values between 40mg / dL and 100mg / dL; HDL size between 7.0 nm and 9.5 nm; CEC between 8% and 20%; antioxidant activity between 0 and 100%. For the remainder of the regression analyzes, we used the entire sample size of the sub-sample of 99 individuals. Two-sided p≤0.05 were considered to v ^®^indicate statistical significance. Analyses were performed using IBM SPSS ersion 21.0 statistical software. Scatter plot was performed using GraphPad Prism version 7.0 for Mac, GraphPad Software, La Jolla California USA, www.graphpad.com”. Polinomial splines were used to evaluate the relation between BMI or carotid IMT vs CEC or Antioxidant activity or a compound variable, after transforming CEC and Antioxidant activity into positive z-scores. The compound variable (Z-Efflux + Z-Antioxidant activity) was defined by the sum of z-scores for CEC and antioxidant activity. In order to avoid overfitting, we excluded extreme values at x-axis (<5th percentile and >95%percentile) and smoothing parameters for polinomial derivation (degrees of freedom [df] and lambda) were defined after cross validation. Spline curves were performed in R software, version 3.2.1 (R Foundation for Statistical Computing). We used MATLAB^®^ (version 4.10, The MathWorks, Inc, Apple Hill Drive Natick, MA) to fit a 3D curve using second polynomial least square surface.

## 3. Results

### 3.1. Overall study participants characteristics

The baseline characteristics of participants are shown in Table 1 stratified by BMI status (n=899). The female gender prevailed among the participants and BMI values were associated with waist circumference, increased cIMT and impaired metabolic parameters; *i.e*. lower HDL-C, Apo-AI and HOMA2%S, as well as higher triglycerides, insulin and fasting glucose. Similarly, a stepped increase in LDL-C and total cholesterol was also noted with increasing BMI.

**Table 1.** Clinical characteristics according to BMI groups

|  | BMI categories (kg/m<sup>2</sup>) |  |  | *p-value* |
| --- | --- | --- | --- | --- |
|  | <22 | 22-24.99 | 25-30 |  |
| **Sample size** | 186 | 264 | 449 |  |
| **BMI, kg/m<sup>2</sup>** | 20.6 (1.8) | 23.6 (1.6)<sup>a</sup> | 28 (4.6)<sup>b,c</sup> | <0.0001 |
| **Waist circumference, cm** | 68 (9) | 75 (11)<sup>a</sup> | 94 (19)<sup>b,c</sup> | <0.0001 |
| **Age, yr** | 38 (24) | 45.5 (26)<sup>a</sup> | 54 (16)<sup>b</sup> | 0.003 |
| **Male sex, %** | 43.5 | 44.7 | 39.6 | 0.369 |
| **Hypertension, %** | 28.6 | 31.3 | 28.3 | 0.915 |
| **Glucose, mg/dL** | 81 (12.0) | 86 (14.0)<sup>a</sup> | 105 (62.0)<sup>b,c</sup> | <0.0001 |
| **Insulin, μU/mL** | 3.0 (2.5) | 4.7 (4.9)<sup>a</sup> | 10.2 (8.2)<sup>b,c</sup> | <0.0001 |
| **HOMA2 % $\beta$** | 61.9 (38.2) | 61.3 (49.4) | 64.9 (51.3)<sup>b,c</sup> | 0.008 |
| **HOMA2 %S** | 241.7 (210.3) | 161.2 (183.5) | 68.8 (62.5)<sup>b,c</sup> | <0.0001 |
| **HOMA2-IR** | 0.46 (0.4) | 0.51 (0.5) | 0.66 (0.5)<sup>b,c</sup> | <0.0001 |
| **CRP, mg/dL** | 0.42 (0.9) | 0.70 (1.1) | 0.51 (1.0) | 0.453 |
| **Triglycerides, mg/dL** | 66 (34) | 81 (46)<sup>a</sup> | 118 (87)<sup>b,c</sup> | <0.0001 |
| **LDL-C, mg/dL** | 95 (40) | 104 (34)<sup>a</sup> | 107 (42)<sup>b</sup> | <0.0001 |
| **Total cholesterol, mg/dL** | 175 (53) | 181 (50) | 187 (55)<sup>b,c</sup> | < 0.0001 |
| **ApoA-I, mg/dL** | 148 (53) | 142 (46) | 137 (38)<sup>b,c</sup> | <0.0001 |
| **ApoB, mg/dL** | 72 (23) | 81 (27)<sup>a</sup> | 90 (31)<sup>b</sup> | <0.0001 |
| **HDL-C, mg/dL** | 63 (29) | 52 (30)<sup>a</sup> | 45 (19)<sup>b,c</sup> | <0.0001 |
| **cIMT, mm** | 0.55 (0.20) | 0.60 (0.20)<sup>a</sup> | 0.70 (0.24)<sup>b,c</sup> | < 0.0001 |

All continuous variables are expressed as medians and interquartile ranges, and categorical variables as percentages. Comparisons between groups were analyzed with Jonckheere– Terpstra’s for ordered alternatives with pairwise comparisons, or Chi-Square test for the categorical variables. To convert the values for glucose to millimoles per liter, divide by 18. To convert the values for insulin to picomoles per liter, multiply by 6.945. To convert the values for cholesterol to millimoles per liter, multiply by 0.02586. To convert the values for triglycerides to millimoles per liter, multiply by 0.01129. <sup>a</sup>BMI 22 – 24.9 vs BMI <22; <sup>b</sup>BMI >25 vs 22 – 24.9; <sup>c</sup>BMI > 25 vs BMI < 22. Significant p-value < 0.05.

### 3.2. Association between BMI, HDL-C and cIMT

In the whole studied sample (n=899), multivariable linear regression analyses adjusted for BMI, age, gender, and plasma insulin or HOMA2-IR, confirmed the association between cIMT and HDL-C (β=-0.11; 95%CI=-0.020;0.000; p=0.03) and identified the existence of a significant interaction of BMI upon this association (β=-1.80; p<0.0001). To visualize the pattern for such BMI interaction, 3D surface plots were applied based on second polynomial least square regression (Figure 1). As shown in the Figure 1A, in subjects with increased BMI, reduced HDL-C levels were associated with increased cIMT. In contrast, in those individuals with reduced BMI, no clear association was found between HDL-C and cIMT. In order to provide a deeper assessment of this interaction, a broad spectrum of HDL functions was investigated in a subgroup of these individuals. As it would be expected, an inverse association was found between BMI and HDL-C (Spearman’s rho -0.028, p < 0.0001) and HDL size (Spearman’s rho -0.175, p < 0.0001).

**Figure 1.**
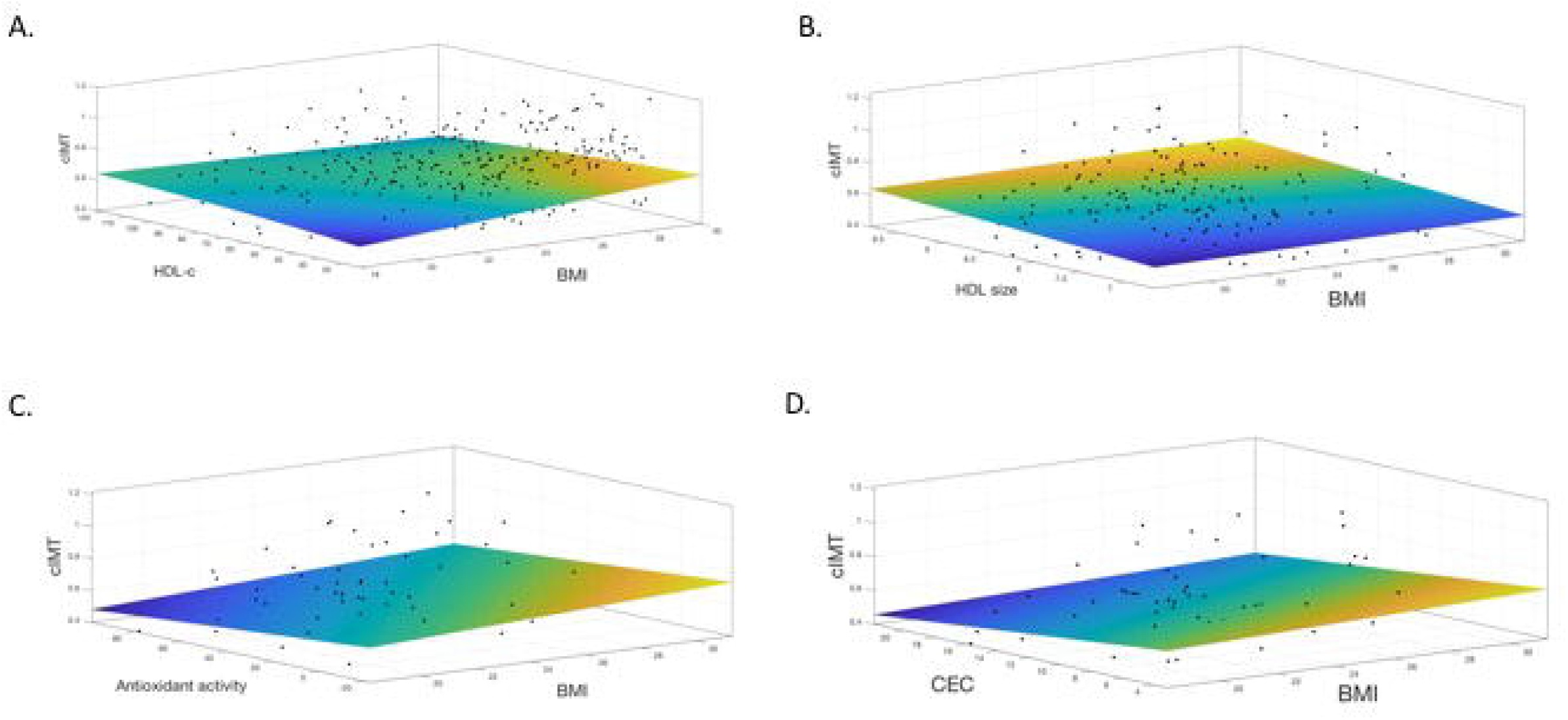
3D surface plot showing the association between BMI, cIMT, and HDL variables. This surface plot displays an image based on the relationship between the BMI and HDL variables as predictors on the x- and y-axes and a continuous surface that represents the cIMT values on the z-axis. The peak on the plot corresponds with the yellow color and the highest value obtained to cIMT using the combination of X and Y that produce the maxima cIMT, which occurs at approximately 0.8mm. The valley corresponds with blue color and the combination of X and Y that produce the minima cIMT. Figure 1A, HDL-C values. Figure 1B, HDL size. Figure 1C, CEC values. Figure 1D, antioxidant activity of HDL. HDL-C, high-density lipoprotein cholesterol in mg/dL. BMI, body mass index in kg/m^2^. CEC is expressed as a percentage of efflux in the sample, normalized to a reference sample. Antioxidant activity is expressed in % as inhibition of LDL oxidation in the presence of each subject’s HDL oxidation.

### 3.3. Association between BMI, HDL functions and cIMT in the subgroup analyses

The subgroup characteristics are shown on Table 2. In the subgroup, we found a stepped decrease of total HDL mass, HDL size, and HDL content in triglyceride, free cholesterol, cholesteryl ester, and phospholipid across BMI categories (Table 3). PON activity adjusted for the total number of HDL particles, *i.e*. PON/HDL-C ratio, increased in parallel with BMI categories. There was no relation between BMI categories and activities of PLTP, LCAT, HL or LPL (Table 3).

**Table 2.**
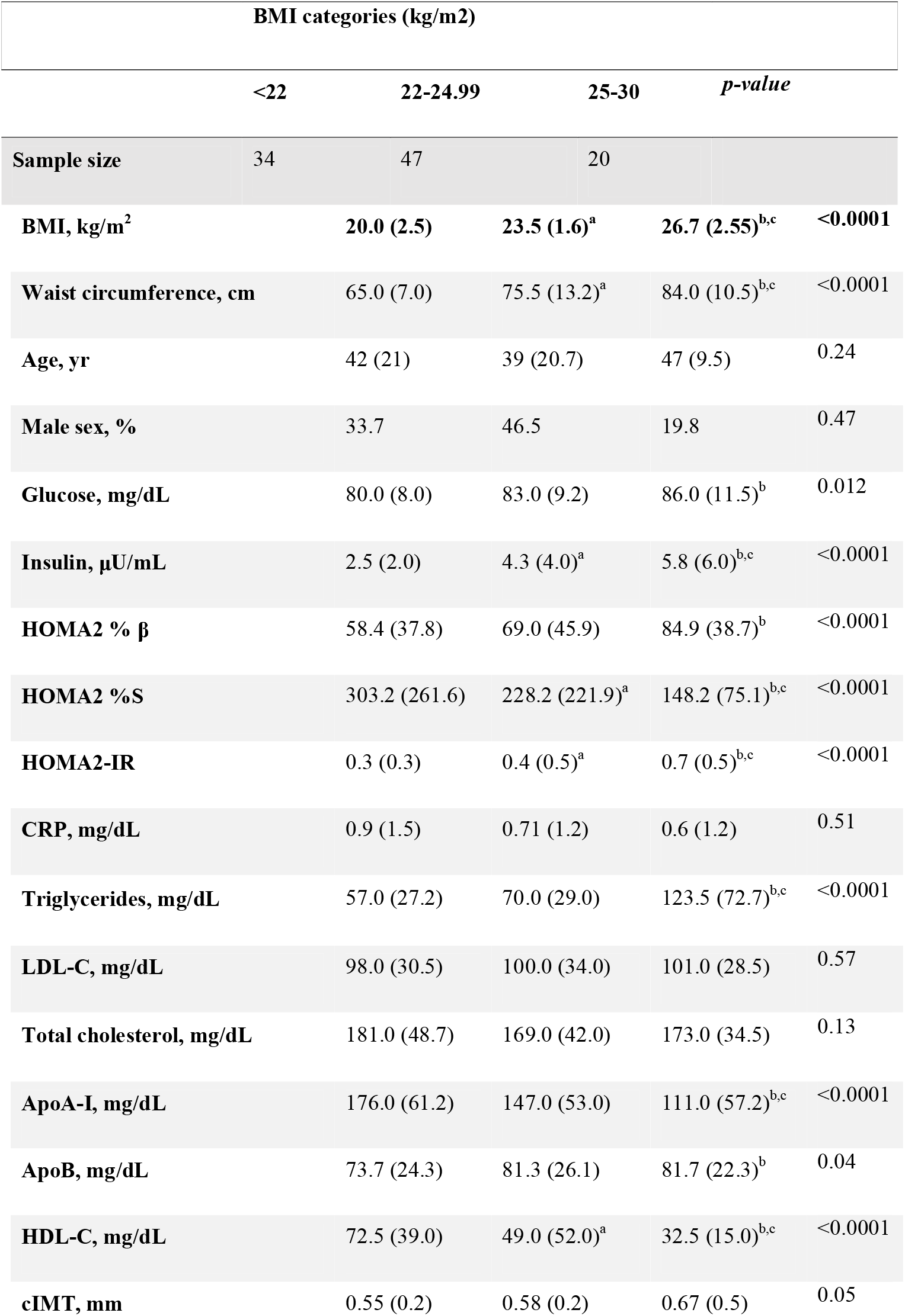
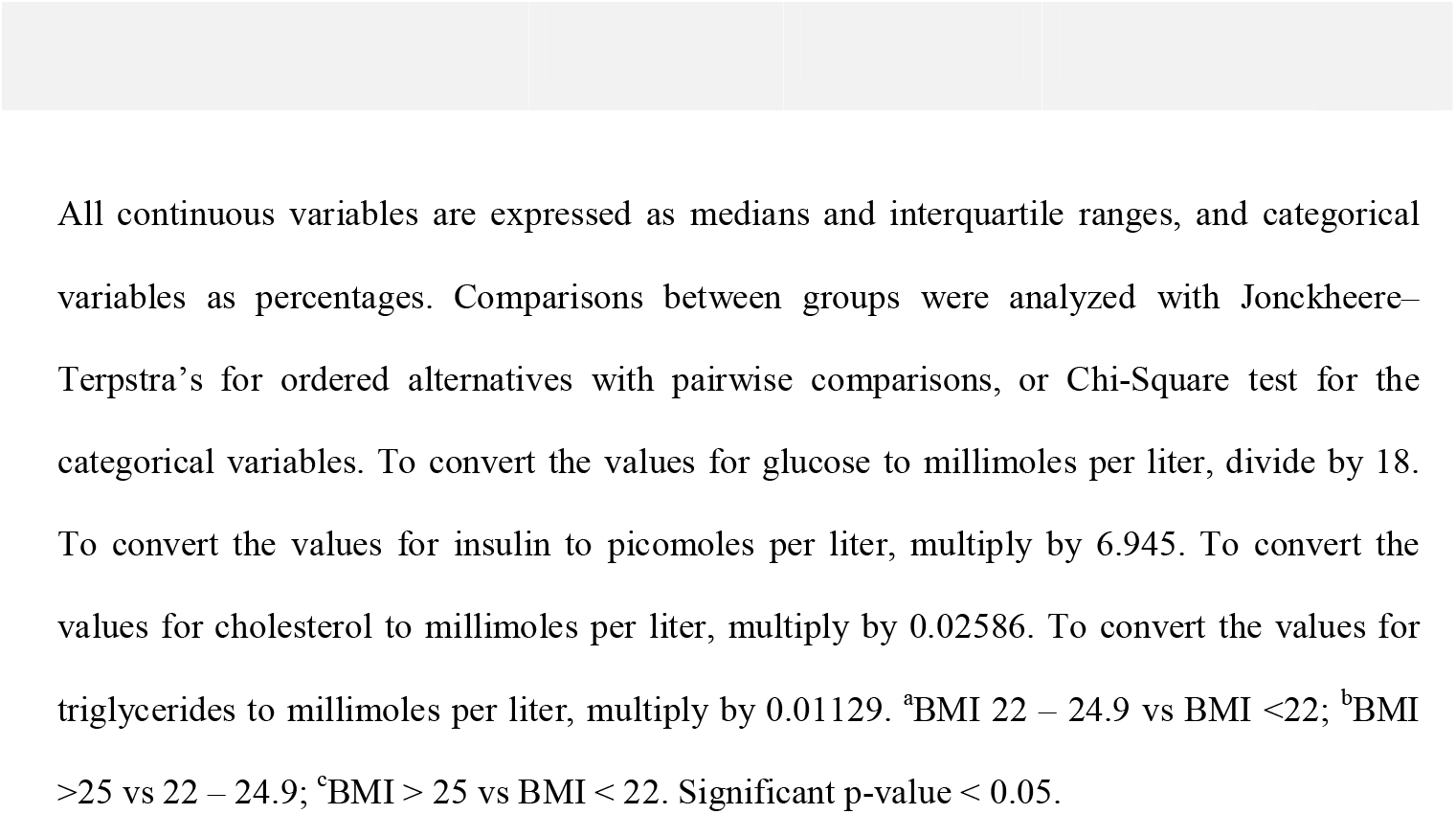
Subgroup clinical characteristics according to BMI groups

**Table 3.**
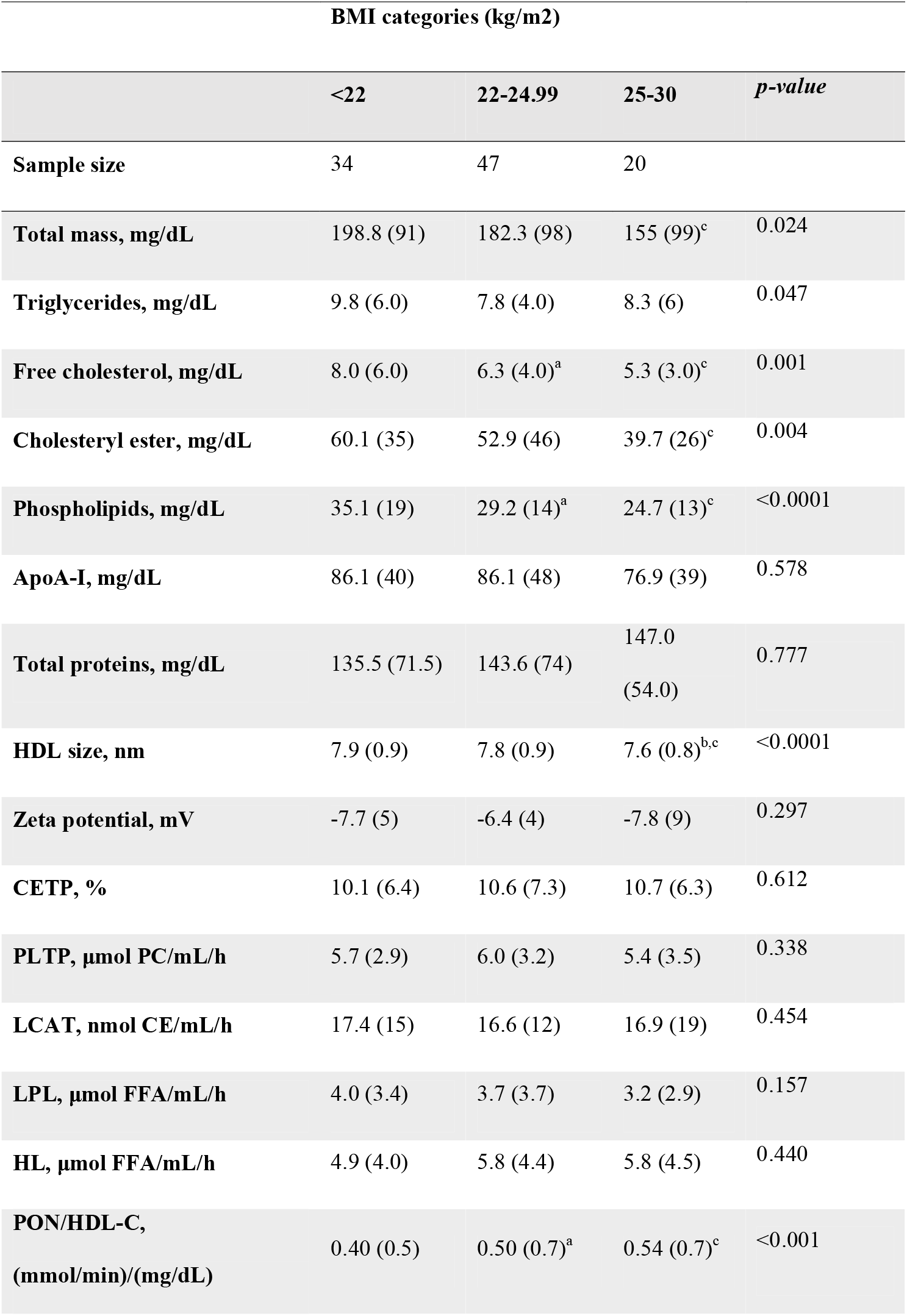
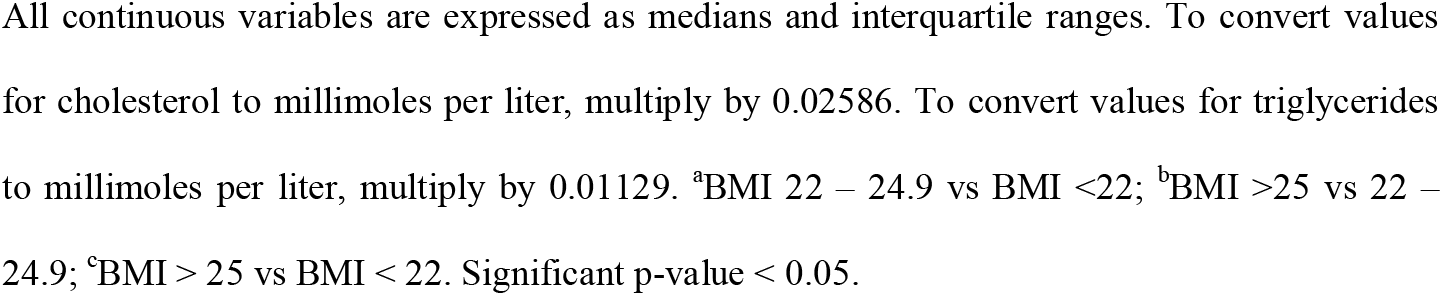
HDL characterization

|  | BMI categories (kg/m<sup>2</sup>) |  |  | *p*-value |
| --- | --- | --- | --- | --- |
|  | <22 | 22-24.99 | 25-30 |  |
| **Sample size** | 34 | 47 | 20 |  |
| **Total mass, mg/dL** | 198.8 (91) | 182.3 (98) | 155 (99)<sup>c</sup> | 0.024 |
| **Triglycerides, mg/dL** | 9.8 (6.0) | 7.8 (4.0) | 8.3 (6) | 0.047 |
| **Free cholesterol, mg/dL** | 8.0 (6.0) | 6.3 (4.0)<sup>a</sup> | 5.3 (3.0)<sup>c</sup> | 0.001 |
| **Cholesteryl ester, mg/dL** | 60.1 (35) | 52.9 (46) | 39.7 (26)<sup>c</sup> | 0.004 |
| **Phospholipids, mg/dL** | 35.1 (19) | 29.2 (14)<sup>a</sup> | 24.7 (13)<sup>c</sup> | <0.0001 |
| **ApoA-I, mg/dL** | 86.1 (40) | 86.1 (48) | 76.9 (39) | 0.578 |
| **Total proteins, mg/dL** | 135.5 (71.5) | 143.6 (74) | 147.0 (54.0) | 0.777 |
| **HDL size, nm** | 7.9 (0.9) | 7.8 (0.9) | 7.6 (0.8)<sup>b,c</sup> | <0.0001 |
| **Zeta potential, mV** | -7.7 (5) | -6.4 (4) | -7.8 (9) | 0.297 |
| **CETP, %** | 10.1 (6.4) | 10.6 (7.3) | 10.7 (6.3) | 0.612 |
| **PLTP, μmol PC/mL/h** | 5.7 (2.9) | 6.0 (3.2) | 5.4 (3.5) | 0.338 |
| **LCAT, nmol CE/mL/h** | 17.4 (15) | 16.6 (12) | 16.9 (19) | 0.454 |
| **LPL, μmol FFA/mL/h** | 4.0 (3.4) | 3.7 (3.7) | 3.2 (2.9) | 0.157 |
| **HL, μmol FFA/mL/h** | 4.9 (4.0) | 5.8 (4.4) | 5.8 (4.5) | 0.440 |
| **PON/HDL-C,**<br>**(mmol/min)/(mg/dL)** | 0.40 (0.5) | 0.50 (0.7)<sup>a</sup> | 0.54 (0.7)<sup>c</sup> | <0.001 |

All continuous variables are expressed as medians and interquartile ranges. To convert values for cholesterol to millimoles per liter, multiply by 0.02586. To convert values for triglycerides to millimoles per liter, multiply by 0.01129. <sup>a</sup>BMI 22 – 24.9 vs BMI <22; <sup>b</sup>BMI >25 vs 22 – 24.9; <sup>c</sup>BMI > 25 vs BMI < 22. Significant p-value < 0.05.

In line with previous studies, as shown in Figure 2A (linear beta -0.38, polynomial R2=0.19, p=0.001; Spearman’s rho -0.32, p<0.001), we reported a progressive decline in CEC as BMI increases. In contrast, we found a progressive increase in HDL antioxidant activity which was proportional and reciprocal as compared with CEC (linear beta +1.38, polynomial R2=0.12, p=0.04) (Figure 2B). Both changes were mainly mediated by the HDL size which is inversely related to weight gain. Based on this assumption, the compound variable of z-transformed efflux plus antioxidant activity should remain approximately constant as HDL size changes due to excess weight (p value 0.75) (Figure 2C). Increasing values of the compound variable were associated lower carotid IMT (linear beta -0.10, polynomial R2=0.26, p<0.001) (Figure 2D). Still, correlation analyses were made between BMI and HDL functions. An inverse association was also found with HDL-mediated platelet inhibition (Spearman’s rho -0.157, p < 0.03). No correlation was found between BMI and HDL anti-inflammatory activity (Spearman’s rho -0.009, p < 0.904) nor between each of the HDL functions.

**Figure 2.**
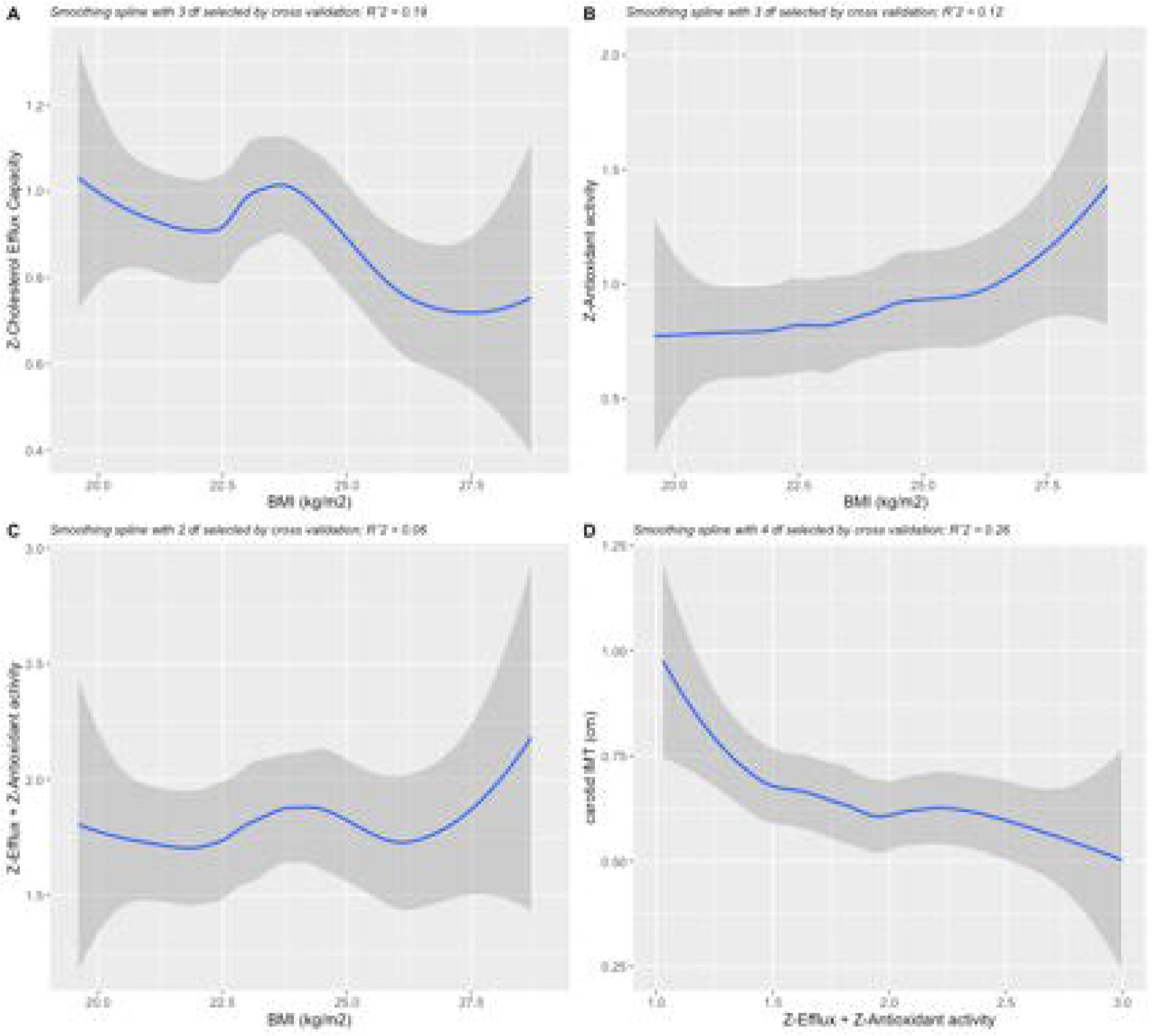
Polynomial splines showing the associations between BMI, cIMT and HDL variables. 2A, CEC according to BMI values. 2B, Antioxidant activity according to BMI values. 2C, compound variable (Z-Efflux + Z-Antioxidant activity) according to BMI values. 2D, cIMT values according to compound variable (Z-Efflux + Z-Antioxidant activity). Gray shading indicates 95% confidence intervals.

Since both the abundance of substrate, *i.e*. triglyceride-rich lipoproteins, and the activities of transport proteins are potentially involved in the effect of excess weight on HDL phenotype, multivariate linear regression models were applied to estimate the influence of intravascular HDL-remodeling proteins, *i.e*. CETP, LPL, HL, LCAT and PLTP on HDL size. CETP (β=-0.12; 95%CI=-0.30;-0.01; p=0.030), HL (β=-0.28; 95%CI=-0.35;-0.12; p<0.0001) and LCAT (β=-0.36; 95%CI=-0.28;-0.13; p<0.0001) were drivers for HDL size. HDL size was also associated with plasma triglycerides (β=-0.17; 95%CI=-0.06;-0.02; p<0.001), HOMA2-IR (β=0.21; 95%CI=0.01;0.42; p=0.001), PON activity/HDL-C (β=-0.33; 95%CI=-0.47;-0.13; p<0.0001) and HDL-mediated antioxidant activity (β=-0.70; 95%CI=-0.81;-0.12; p=0.009). Association was found between HOMA2-IR and HDL-mediated antioxidant activity (β=-2.1; 95%CI=-3.66;-0.60; p=0.007).

In order to estimate the association between HDL functions and cIMT, we used multiple linear regression with adjustments for the following confounders (BMI, age, sex, HDL-c and insulin), which were selected based on univariate significance or clinical relevance (age, sex) (Table 4). We observed a negative association between CEC and cIMT, which remained significant after initial adjustment for the same confounders above, or even after adjustment for HDL size (β=-0.18; 95%CI=-0.138; -0.011; p=0.02), antioxidant activity (β=-0.19; 95%CI=-0.003;0.000; p=0.04) and PON activity/HDL-C (β=-0.16; 95%CI=-0.065;-0.002; p=0.02). The antioxidant activity of HDL was also inversely related to cIMT. Anti-inflammatory activity, PON activity/HDL-C and HDL-mediated platelet inhibition were not associated with cIMT in an adjusted model.

**Table 4.** Linear regression analyses between HDL functions and cIMT

| Independent variable | Unstandardized $\beta$ | Standardized $\beta$ | Lower CI 95% | Upper CI 95% | *p*-value |
| --- | --- | --- | --- | --- | --- |
| Antioxidant activity |  |  |  |  |  |
| Unadjusted |  |  |  |  |  |
| Adjusted<sup>a</sup> | -0.001 | -0.153 | -0.003 | 0.000 | 0.128 |
|  | -0.002 | -0.196 | -0.003 | 0.000 | 0.038 |
| CEC |  |  |  |  |  |
| Unadjusted | -0.016 | -0.260 | -0.028 | -0.004 | 0.009 |
| Adjusted<sup>a</sup> | -0.012 | -0.188 | -0.023 | 0.000 | 0.047 |
| PON/HDL-C |  |  |  |  |  |
| Unadjusted | -0.010 | -0.063 | -0.079 | 0.024 | 0.301 |
| Adjusted<sup>a</sup> | -0.045 | -0.061 | -0.071 | 0.019 | 0.254 |
| Anti-inflammatory activity |  |  |  |  |  |
| Unadjusted | 0.000 | -0.043 | -0.003 | 0.002 | 0.676 |
| Adjusted<sup>a</sup> | 0.000 | -0.004 | -0.002 | 0.002 | 0.963 |
| HDL-mediated platelet inhibition |  |  |  |  |  |
| Unadjusted | 0.002 | 0.164 | 0.000 | 0.004 | 0.120 |
| Adjusted<sup>a</sup> | 0.002 | 0.177 | 0.000 | 0.004 | 0.068 |

<sup>b</sup> Adjusted for age, sex, BMI, HDL-C and insulin. In order to compare effects across different HDL functions we provided the standardized coefficients.

Interactions of BMI were found within the linear associations between cIMT and CEC (p=0.03), and cIMT and antioxidant activity (p=0.04). Although the 95% CI suggests a tendency towards a moderating effect of the BMI on the association between HDL size and cIMT, this interaction did not reach statistical significance (p=0.13) (Fig. 1B). As displayed in Fig 1C, the inverse association between CEC and cIMT seems to increase in parallel with BMI values. In contrast, as shown in Fig 1D, the inverse association between cIMT and antioxidant activity of HDL increases with increasing BMI. In fact, the correlation between cIMT and antioxidant activity is greater in individuals with BMI above the median (23.6 kg/m2) (r=-0.33; p=0.037) than in their counterparts (r=-0.2; p=0.05) while the correlation between CEC and cIMT is equal in both subgroups (above, r=-0.28; p=0.042; below, r=-0.28;p=0.016).

## 4. Discussion

Recent data have shown that the predictive value of HDL-C for estimating the risk of cardiovascular events declines after CVD manifestation[23]; a failure that has been attributed to the increased generation of dysfunctional HDL in a setting of chronic or acute disease. As pointed out in subjects with CVD, we hypothesized that excess weight may influence the relationship between HDL-C and the risk of atherosclerotic disease. After confirming this hypothesis, we moved forward investigating whether more subtle changes in the HDL system could occur in overweight individuals and whether this could justify a change in the association pattern between HDL-C and atherosclerotic disease. Taken together, our findings revealed the following evidence: (i) the pattern of association between HDL-C and cIMT differs according to the presence or absence of excess weight; (ii) increased BMI is associated with the simultaneous change of multiple anti-atherosclerotic functions of HDL, including CEC and antioxidant activity; (iii) the existing association between excess weight and carotid atherosclerotic burden is in part attributable to HDL dysfunction; and (iv) HDL-mediated inhibition of platelet aggregation declines with excess weight.

To the best of our knowledge, only few studies have previously assessed the impact of body weight on HDL function in non-obese individuals and none used a simultaneous assessment of multiple functions. In this clinical setting, CEC has been the main HDL function assessed, which was investigated in interventional [3], case-control[24] and cross-sectional [5] studies. In a large nested case-control study from the EPIC-Norfolk cohort, for example, discrete changes in BMI in the overweight range was inversely correlated with CEC [24]. On this specific aspect, our results are in line with previous studies.

Weight gain promotes a wide range of metabolic changes, among which insulin resistance is the cornerstone for CVD risk. As a result of insulin resistance, for example, there is an increase in the fatty acid efflux and in the activities of CETP and HL leading to a reduction in HDL size, an increase in the ApoA-I catabolic rate and a reduction in CEC[5, 25, 26]. In contrast with the evidence in obese subjects [27-29], our study did not identify changes in CETP and HL activities across BMI categories although their activities were associated with HDL size change. Thus, at this stage of excess weight, the biochemical and size changes in HDL particles is more likely a consequence of increased substrate for HDL particle remodeling, *i.e*. increased plasma concentration of triglyceride-rich lipoproteins.

These changes in the HDL system have traditionally been reported as a component cause for the increased risk of CVD in obese individuals[30]. In those who are overweight but below the obesity threshold, we revealed a similar scenario, *i.e*. insulin resistance inversely relates with overweight, HDL size and CEC. It is interesting to note that these changes occurred even before the threshold for overweight as defined by World Health Organization (WHO). Thus, according to previous translational studies[31] or actuarial analyzes based on observational cohorts[32], our data support the concept that the threshold for pathogenic body mass is probably below the WHO definition.

Although it is clear that there is functional impairment, it is plausible that the change in the HDL phenotype would have an adaptive biological purpose motivated by the chronic increase of oxidative stress and by low-grade systemic inflammation. Similar reduction of HDL size has been reported in individuals who exhibit acute phase response, such as in myocardial infarction[33] and sepsis[34]. In these conditions, the reduction of HDL size is associated with a greater capacity to mitigate the oxidative stress. In agreement with this and consistent with previous studies, HDL size was inversely associated with increased antioxidant and PON activities[35]. Interestingly, the predominance of small HDL (7.3-8.2 nm) in this cohort was associated with an attenuated impact of BMI on cIMT. Hence, overweight-induced remodeling in HDL simultaneously promoted the reduction of an anti-atherosclerotic mechanism, i.e. CEC, and the increase of another, i.e. antioxidant activity, suggesting a biological response of the HDL system to the predominant pro-atherosclerotic stimulus. Besides, the present study shows that estimates of the magnitude of HDL participation in the CVD risk in excess weight are naturally imprecise unless a broad range of anti-atherosclerotic functions is simultaneously investigated.

Obesity is clearly shown to be a prothrombotic state resulting from a combination of increased thrombin generation, platelet hyperactivity and decreased fibrinolysis[36]. Increased platelet reactivity is the result of the interaction between multiple characteristics grouped into obesity, including inflammation, oxidative stress, insulin resistance and adiposopathy with change in adipokine secretion pattern. Our study added to this state of knowledge, a new mechanism of imbalance in platelet activity; inhibition of HDL-mediated platelet aggregation was inversely associated with BMI. This finding should be considered in the set of prothrombotic changes of excess weight and as one of the mechanisms by which there is an increase in the incidence of cardiovascular events in these individuals.

Among the main limitations in the present study lies the fact that it is based on a cross-sectional design. In addition, due to the intensive laborious methodology involved in studying a full breadth of HDL function and metabolic pathways, we may have not been able to maintain the desired statistical power obtained as when the entire studied population was analyzed. In spite of this, it does provide us a unique opportunity to evaluate the association between excess weight and the HDL system early in the spectrum of metabolic derangement and atherogenesis. In fact, this may especially be true given the low rates of insulin resistance and more favorable lipid profile and systemic inflammatory activity of our study’s participants.

In conclusion, our study indicates that higher BMI, even in non-obese, non-diabetic individuals, is associated with decreased function of several protective activities of HDL. In individuals with increased BMI, HDL dysfunction is directly associated with increased atherosclerotic burden. Antioxidant activity is found to be an exception increasing with BMI and it partially attenuated the atherosclerotic burden.

## Data Availability

The datasets generated during and/or analysed during the current study are available from the corresponding author on reasonable request.

## Abbreviations

ALT: alanine aminotransferase
ApoA-I: apolipoprotein A-I
AST: aspartate aminotransferase
CE: cholesteryl ester
CEC: Cholesterol efflux capacity
CETP: cholesteryl ester transfer protein
cIMT: carotid intima-media thickness
FC: free cholesterol
HL: hepatic lipase
HOMA2 % β: Homeostasis model assessment 2 of beta-cell function
HOMA2-IR: Homeostasis model assessment 2 of insulin resistance
HOMA2 %S: Homeostasis model assessment 2 of insulin sensitivity
HUVEC: human umbilical vein endothelial cells
LCAT: Lecithin–cholesterol acyltransferase activity
PL: phospholipidis
LPL: Lipoprotein lipase
PON: paraoxonase
TC: total cholesterol
PLTP: phospholipids transfer protein
VCAM-1: vascular cell adhesion molecule-1

## Author contribution statement

Jose Carlos de Lima-Junior drafted the manuscript and participated on the analysis of the data, Vitor W.M. Virginio participated on the experiments, Filipe A. Moura participated on the drafting of the manuscript and analysis of the data, Adriana Bertolami participated on the experiments, Marcelo Bertolami participated critically reviewing the manuscript, Wilson Nadruz participated critically reviewing the manuscript, Eliana Cota de Faria participated on the experiments and analysis of the data, Ilaria Zanotti participated critically reviewing the manuscript, Luiz Sergio F. de Carvalho participated on the analysis of the data and critically reviewing the manuscript, Andrei C Sposito participated on the analysis of the data and critically reviewing the manuscript

## Acknowledgements

We thank Natalia B. Panzoldo from the University of Campinas for technical assistance in HDL functions experiments.

